# PlasmoCount 2.0: Rapid Multi-Species Malaria Parasite Detection Using Deep Learning

**DOI:** 10.1101/2025.05.05.25326942

**Authors:** Frank Weate, Yunchuan Li, David Novotny, Erik Meijering, Jake Baum

**Affiliations:** School of Biomedical Sciences, University of New South Wales, Sydney, Australia; School of Computer Science and Engineering, University of New South Wales, Sydney, Australia; Department of Life Sciences, Imperial College London, London SW7 2AZ, UK

**Keywords:** Artificial intelligence, Giemsa stain, light microscopy, residual neural networks (ResNets), Vision transformer (ViT), ConvNeXt, You Only Look Once (YOLO, convolutional neural network (CNN)

## Abstract

Visual examination of Giemsa-stained red blood cell smears is the gold-standard for identification of malaria parasite infection. Despite its wide usage, however, smear counting is time consuming, and the storage of slides has limited archiving or referencing capacity. Towards automation of smear counting and to support digital archiving, we previously developed a deep-learning application, called PlasmoCount, that provides accurate, model-assisted counting of intracellular parasites using convolutional neural networks. PlasmoCount was specifically designed for use with the human malaria parasite *Plasmodium falciparum*. Here, we overhaul the platform, improving its versatility and robustness. Principally, we have updated the tool so that it is capable of detecting blood-stage infections from multiple species of human-infective and rodent-infective *Plasmodium* parasites and have adapted it for use with both 40x and 100x objective magnifications. These augmentations broaden the distribution of input data our model can accommodate whilst maintaining its high classification accuracy (99.8%), along with modestly improving its precision with cell detection. We see significant prediction improvements on out-of-domain data, showing the adaptability of the model for real-world applications. In addition, we have substantially reduced the processing time of PlasmoCount by replacing Faster R-CNN, the original object detection model, with YOLOv8 and using batch inference for classification - modifications that reduce the latency by 90%, processing a single image in under 3 seconds (reduced from 40). Finally, we provide an offline, on-device version of the standardised framework, now referred to as PlasmoCount 2.0, which is compatible with most smartphones, including iOS and Android operating systems. PlasmoCount 2.0 markedly improves the time taken for malaria parasite smear-based detection and provides a reproducible means to assess parasite infections in laboratory settings as well as providing a roadmap for future application of the platform in clinical or field diagnosis.

## INTRODUCTION

Malaria is an infectious disease caused by infection with *Plasmodium* spp. parasites transmitted by a mosquito bite. It is a disease of major global significance, responsible for an estimated 597,000 deaths in 2023 with the majority of deaths in children under five in sub-Saharan Africa^1^. Light microscopy inspection of thin and thick blood smears, identifying and counting infected red blood cells (RBCs) stained with Giemsa, remains the gold standard for diagnosing malaria. This method provides additional valuable information that can guide treatment decisions such as the parasitaemia (percentage of infected cells), parasite lifecycle stage and can aid in determining the *Plasmodium* species responsible. Giemsa-smear inspection is also an important complementary diagnostic approach that can address evolving resistance of some parasites to lateral-flow based rapid diagnostic tests^2^. Moreover, microscopy is also widely used in laboratory research that rely on *in vitro* (culture-based) and *in vivo* cultivation (rodent models) of malaria parasites, where an accurate parasitaemia prediction is vital for day-to-day scientific programs. A process that increased the efficiency and accuracy of parasitaemia prediction would greatly aid research aiming to develop new drugs and vaccines for malaria and could assist in both infection surveillance and population health efforts towards regional elimination/eradication.

We previously developed a deep learning platform called PlasmoCount^3^, designed to replace the time-consuming and inconsistent nature of manual light microscopy inspection of blood smears. The initial model, trained on a large dataset of slide image data, was developed to target the most virulent human malaria parasite, *Plasmodium falciparum.* PlasmoCount was developed as a three-stage neural network pipeline for the detection and classification of malaria-infected cells in Giemsa-stained images. The first stage made use of advances in object detection models, enabling rapid detection of all RBCs regardless of their infection status. The second stage classified each RBC as infected or uninfected, with a third stage predicting the asexual (replicative) life cycle stage (termed rings, trophozoites and schizonts) of the parasite within the infected RBC.

The final multi-stage model was adapted to an online end-to-end web application, enabling a complete functional tool for researchers to upload images and receive results in an efficient and tractable way. The primary focus of PlasmoCount was to deliver an intuitive and frictionless solution to its users, ensuring that the model could make a significant impact in both research and, with refinement, a potential application in clinical diagnostic environments.

PlasmoCount is not the only platform to explore the application of Machine Learning (ML) methodology to classification and detection of malaria-infected RBC smears. Originally statistical methods such as thresholding, clustering, decision trees and morphological operators were used for this task^4–8^, these models were later surpassed by deep learning methods in both accuracy and robustness^9,10^. Some deep learning approaches have enabled classification on single-cell images^9,11–13^, images in which a single RBC makes up the entirety of the image. Such approaches usually involve training a single convolutional neural network (CNN) classifier to predict the likelihood of a cell, once isolated, as infected or not. These solutions rely on traditional ML algorithms to extract each cell from an image, such as watershed segmentation^14^. Other methods, including the original PlasmoCount platform, focus instead on using deep learning methods to detect infected cells within an entire microscope’s field-of-view^3,10,15^. This is an innately harder task as each microscope field-of-view image contains much more information than images of a single cell. Such deep-learning approaches have shown that partitioning the task into an ensemble of models provides superior performance over sole reliance on a single model, due to the significant class imbalance between infected and uninfected cells.

Beyond model choice, other variables also impact the utility of any platform designed for detection and differentiation of malaria parasite infection of RBCs. One key variable is magnification of images. Previous work^17,18^ has shown that it is possible to use images from both 40x and 100x objectives during training, demonstrating that deep learning techniques can perform just as accurately at 40x as they do at 100x. This may aid in accelerating the pace at which parasite prediction occurs simply by virtue of the fact that 40x magnification objective, with its larger field of view, can detect 525% more cells per image than 100x. A second variable is species specificity.

Different species of *Plasmodium* parasite can exhibit significant morphological differences from one another. Given this morphological variance, a classifier trained on one species will often fail to predict well across other *Plasmodium* spp. Whilst there is some research into CNN image classifiers that can classify parasites by species^16^, there is no research into how the inclusion of distinct *Plasmodium* spp. when training the model affects a binary classifier’s generalisability and accuracy. A broadly useful model would benefit by being able to work agnostically with any species. Finally, any robust model needs to work across a variety of laboratory settings (varying in image quality, staining, microscopy platform etc.). Basing a model on a single dataset of uniform quality and then testing from the same distribution of data as the training dataset often leads to overly optimistic results.

A final consideration is user experience. Several groups provide an end-to-end service, such as the mobile application Malaria Screener^19^, or the Windows and Mac OS Application for rodent malaria, Malaria Screener R^18^. While these applications are valuable, both use a narrow distribution of images during training, potentially leading to high error rates on ‘real-world’ out-of-domain data.

There remains a need for versatile solutions that can predict accurately and robustly on human and non-human malaria parasite infections and successfully navigate the wide variance in data quality that is required for most use cases.

Here we set out to address each of these challenges, augmenting the training data to optimise model prediction across a variety of magnifications. In addition, we aimed to see how a single CNN classifier trained on multiple species compares in accuracy to one trained on a single species.

Finally, to advance broad utility of our platform, as both a laboratory tool and potential clinical diagnostic aid, we have developed a smart-phone compatible platform (iOS and Android) that provides radically boosted speed, and enhanced user-friendliness all with the aim of making the tool as useful as possible for its intended users in the laboratory or clinic. Our interactive system works well across seen and unseen *Plasmodium* species and not only provides model predictions but also allows users to provide their own corrections (through cell editing) creating a positive feedback loop where diverse user input data is collected to refine and improve model accuracy and robustness.

## METHODS

### Giemsa Smear Image Datasets

The original PlasmoCount (version 1.0) used classification and regression models trained on Giemsa smear images acquired from multiple laboratory strains of *P. falciparum* (3D7, NF54, Dd2 and D10, <u>dx.doi.org/10.17632/j55fyhtxn4.1</u>), whilst the object detection model trained both on this primary *P. falciparum* dataset and a secondary *P. vivax* dataset ^20^ from the Broad Bioimage library (<u>bbbc.broadinstitute.org/BBBC041</u>). Here, we added more *P. falciparum* smear images from the National Library of Medicine^21^ and developed a new rodent malaria dataset of Giemsa smears for a total of 2,936 field-of-view images all taken at 100x magnification. This amounts to 286,363 cells across five separate species of malaria. Species used included: the human infective *P. falciparum* and *P. vivax* and rodent malaria parasites *P. berghei*, *P. chabaudi* and *P. yoelii* (Table 1).

**Table 1.** The image and cell count of each *Plasmodium* species from our dataset of Giemsa smear images. w*_u_* and *w_p_* are the sampling weights given to the uninfected and parasitised cells in the dataset to sample both infection and species evenly, preventing model overfitting to a particular species or cell type during training.

| Species | Image Count | Cell Count | $w_u$ | $w_p$ |
| --- | --- | --- | --- | --- |
| *P. falciparum* | 463 | 70,277 | 0.0113 | 0.2137 |
| *P. vivax* | 1,267 | 81,998 | 0.0095 | 0.2726 |
| *P. berghei* | 289 | 41,382 | 0.0196 | 0.2526 |
| *P. yoelii* | 593 | 48,264 | 0.0203 | 0.0667 |
| *P. chabaudi* | 389 | 44,442 | 0.0198 | 0.1139 |
| Total | 2,936 | 286,363 | 0.0805 | 0.9195 |

To validate model utility on both seen and unseen parasite species we also used a validation dataset of 164 *P. knowlesi* and *P. cynomolgi* images (a kind gift from T. Annoura & T. Araki, National Institute of Infectious Diseases (NIID), Tokyo, Japan), which are simian malaria parasite species associated with zoonotic malaria in both Southeast Asia and South America.

### Model Pipeline

PlasmoCount uses a 3-stage model pipeline consisting of an object detection model that outputs the predicted bounding box coordinates for all the RBCs in a micrograph. These cells are then cropped and passed as input to the binary classification model that is tasked with predicting whether the cell is infected. Finally, the infected cell images are fed into a regression model which predicts the developmental stage of the parasite, a summary of the original and new workflow is detailed in Figure 1.

**Figure 1.**
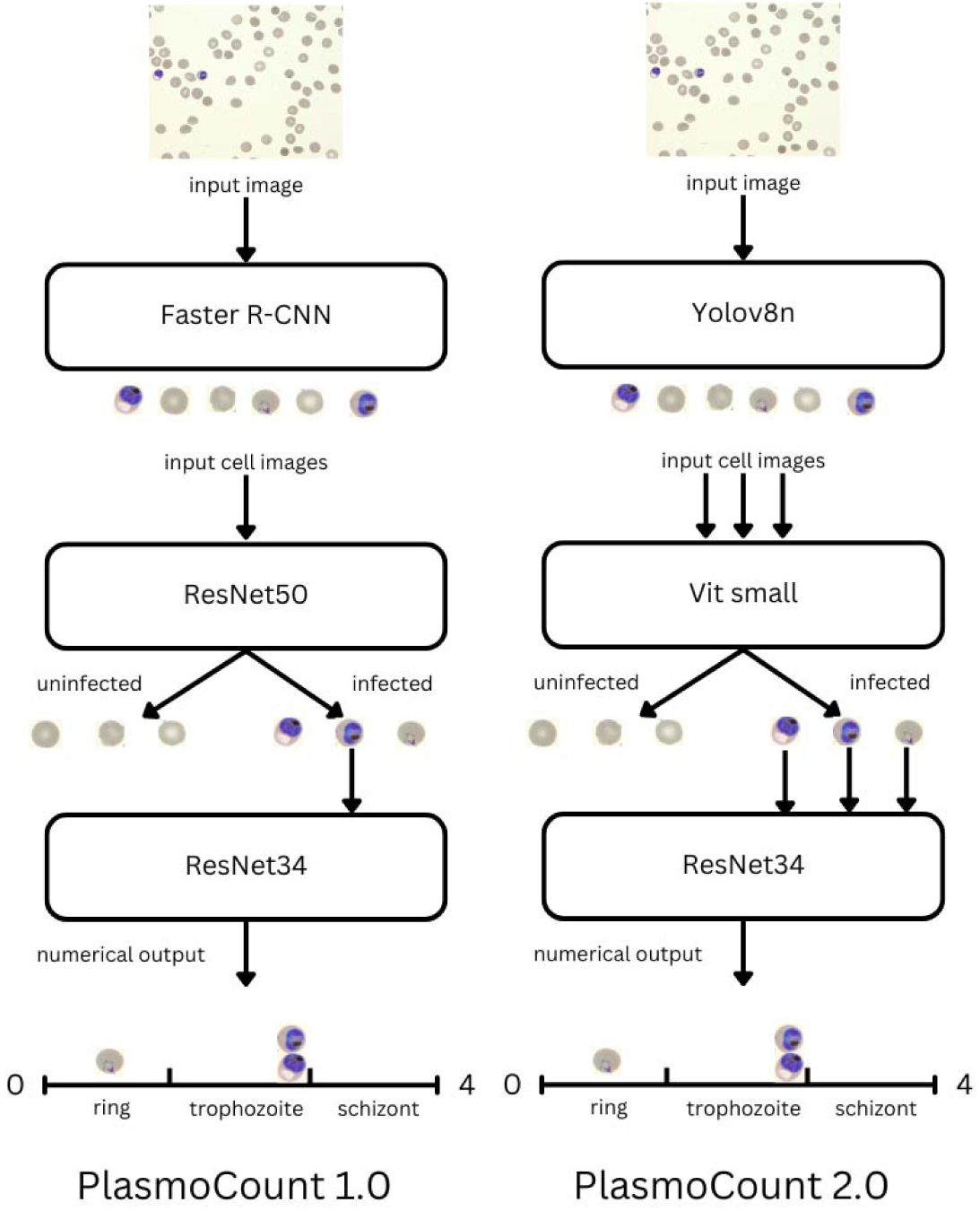
Schematic showing side-by-side comparison of old and new model pipelines for PlasmoCount. Object detection takes in an image of a *Plasmodium* parasite smear slide, stained with Giemsa stain, as input and outputs all detected RBCs. Classification takes cropped cell images of these cells and outputs whether they are infected or not. All infected cells are then handed to the lifecycle model that predicts developmental stage. Multiple arrows in PlasmoCount 2.0 represent inference time batching, significantly reducing prediction time through better utilising CPU hardware.

In developing a new model, we applied the same data augmentation techniques (dihedral transformations, rotation, flipping, and random lighting changes) as used in PlasmoCount 1.0, along with addition augmentations (blurring in the classification models and blurring, mosaic, and scaling in the object detection models). These techniques enhance the robustness of the models with realistic virtual examples. Additionally, noise was added to ground truth coordinates of RBCs to mimic the sub-optimal outputs of the RBC bounding boxes passed to the classification layer.

### Detection Models

The PlasmoCount 1.0 object detection model (Faster R-CNN with FPN, 2017) has been superseded by many recent advances in the field of object detection. Alternative models to Faster R-CNN were surveyed to find one with optimal precision, recall scores and model inference time (to aid running on lower end CPUs in production). We fine-tuned seven models, four variants of You Only Look Once (YOLO) (YOLOv8n, YOLOv8s, YOLOv8m, YOLOv8L), and three transformer architectures (DETR, Deformable DETR, DETA) on our custom dataset. These transformer object models were considered due to their impressive performance on the COCO (Common Objects in Context) dataset - a large-scale image dataset used for object detection, segmentation, and captioning tasks, containing over 330,000 images with detailed annotations for 80 object categories. The three models considered are all based on the DETR (DEtection TRansformer) architecture: DETR, ResNet-50 backbone; Deformable DETR, ResNet-50 backbone; and DeTA Swin Large. Data augmentation strategies were applied equally across all three models. We also analysed another model family, YOLOv8, given its lightweight architecture and extensive use in cell detection tasks. We tested four variants of the YOLOv8 model (nano, small, medium, and large) differing in size, complexity, and general performance. A YOLOv8 training package, ultralytics (www.ultralytics.com), provides additional out of the box data augmentation techniques. As well as random vertical, horizontal flipping, colour changes and blurring, which are all used in the Faster R-CNN model, the package also provided mosaic, random erasing and translating, which were all used in training.

For magnification testing, we also customized the pre-existing scale augmentation from a normal distribution over a scale factor of 1.0 (100x), to a bimodal distribution with one peak at scale factor of 1.0 (100x) and the other peak at scale factor of 0.4 (40x), facilitating sufficient examples of cells to explore the question of model performance with 100x versus 40x magnification. For all models, we set an Intersection over Union (IoU) threshold of 0.5.

### Classification Models

All classification models tested were also pre-trained on ImageNet^23^. A weighted sampler with the dual effect of nullifying class imbalance by oversampling the fewer infected cells in the dataset and sampling between species datasets equally was employed to prevent over-fitting to the common examples in the dataset (see Table 1 for each sample weight). To accurately explore the variety of available classification models, we fine-tuned six different models (ResNet-50, EfficientNet, ConvNeXt Tiny, ConvNeXt, ViT and Densenet121) on the multi-species dataset (n=286,363), and compared this to training the model on species-specific datasets (size given in Table 1). 5-fold cross-validation was used on all tests to evaluate each model’s performance with a higher confidence. The performance of each finetuned model was compared to inform a candidate model choice for final inclusion in the classification layer of the pipeline.

### Multi Magnification Inference

To assess model performance in detecting cells through the microscope with different objective magnifications, 20 images of *Plasmodium*-infected blood smears were captured (Nikon Ds-Ri2 colour camera) using a 100x objective (Plan Apo 100X / NA 1.4 Oil Immersion) or 40x objective (Plan Fluor 40X / NA 0.75) centred on the same field of view. To control for the differences in features between the 40x and 100x (lighting, color, blur), 40x image were also cropped to encompass the exact field of view of the respective 100x image, providing three inputs (100x, 40x and 40x cropped) to compare the output of the model pipeline (YOLOv8n with ConvNeXt-tiny).

### Model Delivery Platforms

With network and cloud loading speeds becoming a limiting factor, we overhauled the original cloud-based architecture working on Android and iOS. Model inference now happens directly on the mobile device, improving the reliability and speed of predictions. Both the Android and iOS versions developed used Flutter (flutter.dev/multi-platform/mobile) to create the UI. The models are converted to PyTorch lightning (github.com/Lightning-AI/pytorch-lightning) and CoreML (developer.apple.com/documentation/coreml/) formats for Android and iOS compatibility, respectively.

### Updated User Experience (UX)

To recapitulate the high UX standard demonstrated in the original platform, PlasmoCount 2.0 was developed to include original data displays (pie charts, histogram, image visualisation) but with added features of cell editing, a progress bar, and error messages. The new cell editing feature allows the microscopist to re-annotate model predictions, even other users’ images. These images, along with their annotations, are stored on the cloud for easy centralised access. This provides PlasmoCount with the potential for a content-rich dataset consisting of extremely diverse images.

## RESULTS AND DISCUSSION

Giemsa staining and microscopy observation of malaria parasite infected blood smears is routine both in laboratory study of human and rodent malarias as well as field-based diagnosis in many resource-poor countries. To better enable the speed of parasite counting, data archiving and standardization of Giemsa smear visualization, we previously developed PlasmoCount (version 1.0), a machine learning tool that provided a cloud based three stage model pipeline—accessible through a web browser—to identify cells, count parasitaemia and classify according to blood stages of development from uploaded images. The original model was trained solely on the most virulent parasite species affecting humans *Plasmodium falciparum*. Whilst accurate and having clear utility, the original platform took several seconds to process images and required internet access. To increase the usability of the platform, we sought to address three key issues in the platform: speed, species generalizability, and ease of use through a smartphone.

### Model Optimization: YOLO Outperforms Transformer Models at Cell Detection

PlasmoCount 1.0 used an object detection model (Faster R-CNN with FPN, 2017) that has since been superseded by many recent advances in the field of object detection. We therefore compared the original Faster R-CNN architecture with four alternative models, three based on the DETR (DEtection TRansformer) architecture and a fourth You Only Look Once (YOLO) model with a lightweight architecture: DETR, ResNet-50 backbone; Deformable DETR, ResNet-50 backbone; DeTA Swin Large and You Only Look Once (YOLO)v8. As demonstrated in **Table 2**, testing of each model demonstrated that substantial improvements in speed could be achieved through implementation of YOLOv8. This model outperformed the other image detection models in our test set, with regards to both precision and speed, reducing processing time by over 90% and demonstrating superior accuracy to PlasmoCount’s original model, Faster R-CNN-FPN (**Table 2**). In addition, we tested four variant YOLOv8 models, nano, small, medium, and large, differing in size, complexity, and general performance. Against expectations, variant models of YOLOv8 had no impact on the model precision, indicating that the additional complexity of the larger YOLO models does not lend itself to a better representation of an RBC. Furthermore, we noted that a transformer models’ high precision on the COCO dataset does not translate to a comparatively higher precision on our RBC dataset.

**Table 2.** Comparison of several object detection models AP_50_ on our dataset and COCO. **Finetuned YOLO models show clear performance benefits over finetuned transformer models on our dataset in both size and accuracy**.

| *Model* | *AP<sub>50</sub> fine-tuned on our dataset* | *AP<sub>50</sub> on COCO* | *Model Parameters (M)* |
| --- | --- | --- | --- |
| *DETR (ResNet-50)* | 87.0 | 63.1 | 41.6 |
| *Deformable DETR (ResNet-50)* | 91.1 | 65.2 | 40.2 |
| *DETA Swin Large* | 88.7 | 69.0 | 219 |
| *Faster R-CNN-FPN* | 98.5 | 58.8 | 41.7 |
| *YOLOv8n* | **99.1** | 52.6 | **3.2** |
| *YOLOv8s* | 99.0 | 61.8 | 11.2 |
| *YOLOv8m* | 99.0 | 67.2 | 25.9 |
| *YOLOv8L* | 99.1 | **69.8** | 43.7 |

Transformer models are generally considered state of the art in many machine learning tasks including object detection in the COCO dataset. Their poor performance on our RBC dataset (**Table 2**) highlights some of the limitations in attention-based models. These architectures rely on an encoder-decoder transformer, in which a larger number of objects lead to more complex scene representation. This imposes greater demands on the decoder, which may struggle with effective object disentanglement and query resolution. Transformer models generally also require long training times and large datasets. While it was possible to train these models for longer with more data augmentations and potentially see a better performance, the performance of the YOLOv8 architecture offers clear benefits as a more convenient, faster, and simpler solution for our pipeline. Finally, a feature of the transformer object detection architecture, which makes it so powerful over CNNs, is global attention for long-range dependencies. The encoder-decoder architecture allows DETR to reason about the relationships between objects and the global image context. This attribute of the architecture makes it perform exceptionally on datasets like COCO, where objects are present in a contextually meaningful scene. However, in the context of Giemsa smears, this does not provide any additional advantages, where the distribution of cells is pseudorandom and uniform. Given the performance of YOLOv8, we selected it as a much faster and more accurate alternative model for object detection in PlasmoCount 2.0.

### Scale Augmentation Successfully Mimics 40x Magnification in Training

We next explored whether PlasmoCount, originally designed for only 100x oil immersion objective imaging, could work at other magnifications either directly or by applying a scaling function to low magnification, in our case 40x objective imaging. As shown in **Table 3**, comparing ”40x (scale)” to “40x” uncorrected images, we see that ”40x scale” captures a similar number of cells to the ”100x” and ”40x (cropped)” images. This demonstrates that the scale data augmentation technique allows the model to learn a very similar representation of images captured with a 40x objective as it does with images captured with a 100x objective and the ”100x” control.

**Table 3.** Model performance on difference capture techniques on the same field of view. “40x (cropped)” is the 40x micrograph cropped to make it artificially appear as 100x, which is then passed to the model. In “40x (scale)” the image is passed to the model at 40x, but the object detection model was trained on a portion of zoomed out 100x images.

| *Magnification* | *Predicted Cell Count* | *Predicted Infected* | *Parasitaemia* |
| --- | --- | --- | --- |
| 100x | 1136 | 108 | 9.51% |
| 40x | 889 | 83 | 9.33% |
| 40x (cropped) | 1122 | 107 | 9.54% |
| 40x (scale) | 1120 | 106 | 9.46% |

Implementing scale augmentation whilst training the object detection model dramatically increased the accuracy of the model on 40x slide-images. When considering the YOLO architecture, particularly the backbone (which generates the feature map of the input image), it is evident that without scale augmentation to introduce sufficient variation in cell size, the model does not incur a loss penalty for missing 40x-sized cells. Of note, just as much deviation is seen with 100x in the ”40x cropped” control as we see in the ”40x scale”. This implies that the difference in lighting, lens and image resolution has a larger impact on cell count than magnification differences. Said another way, this demonstrates that despite the obvious loss of information at lower magnifications, the raw images contain enough core critical features to maintain the performance of models used. Having a model pipeline that can successfully predict 40x magnified images will drastically improve any platforms utility for broad-application malaria detection. Furthermore, assuming a constant cell density, 40x magnification will capture 6.25 as many cells as 100x, saving considerable time to achieve a sufficient sample size.

### Evaluating Speed and Accuracy Trade-Offs in Classification

Having established an improved object detection model that worked across magnifications, we next sought to address potential improvements in image classification (namely infected versus uninfected). In recent years, image classification models have experienced a boon in design improvements in both convolutional and transformer architectures. **Figure 3** shows the speed and error rate of the different models tested here. ResNet-50 is the classification architecture that was incorporated into the original PlasmoCount’s model pipeline. Assessing both error rate and inference time, it is clear that numerous newer models provided promising potential improvements. Of all models tested, ViT-small was both the fastest and had the lowest error rate, the percentage of false predictions in the set of total predictions. For this reason, we selected to incorporate ViT-small as PlasmoCount’s new classification model.

**Figure 3.**
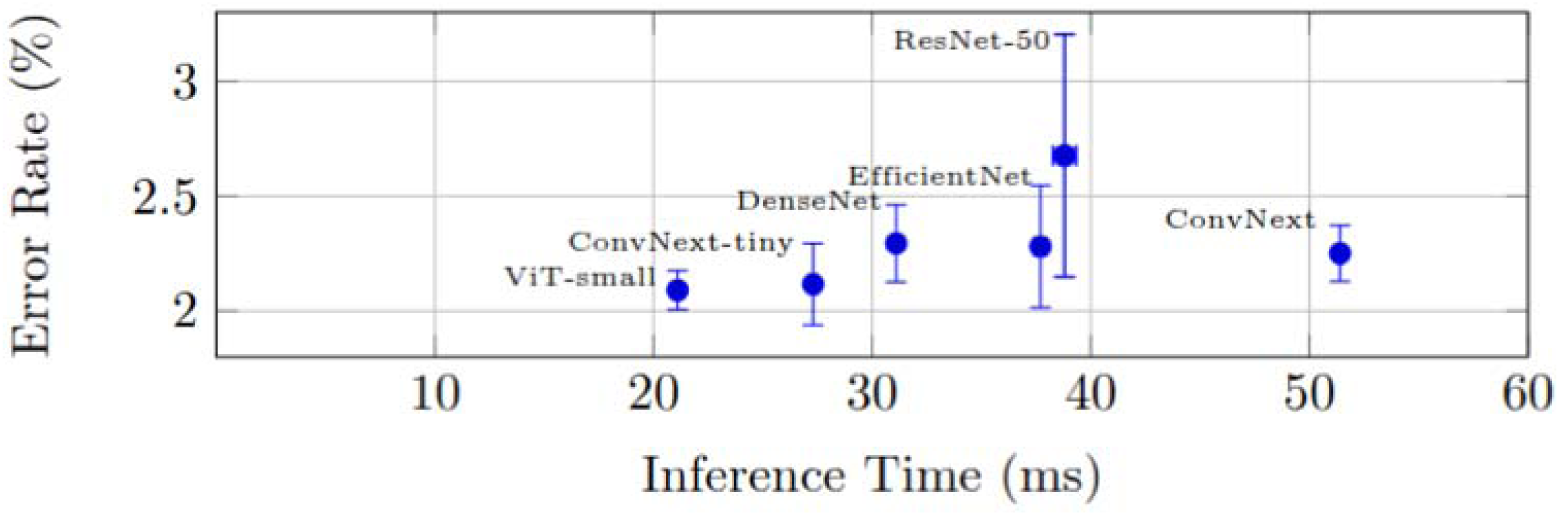
The average error rate observed from 5-fold cross validation in different cell classification (second layer) models. These models were all finetuned on our single cell dataset containing all five species. ViT-small provides both the lowest error rate and inference time, an improvement over ResNet-50 (PlasmoCount 1.0 model).

### Lifecycle stage model: ResNet-34

Due to the small quantity of annotated lifecycle cells (n=2377), it is important to select a model that does not overfit to the training data. Attention models such as ViT, typically require large training datasets to properly fine-tune the complex self-attention weights. ResNet-34, however, is a suitable model as it is shallow and small, and therefore it handles overfitting very well. For this reason, we kept the lifecycle stage model untouched between PlasmoCount 1.0 and 2.0.

### Fine-tuned Classification Models Trained on Multiple Species Perform as well with Single Species

Given rodent species of malaria are crucial for the development of preclinical therapeutics and vaccines^18^, we sought to test whether training on single *Plasmodium* species datasets versus combined *Plasmodia* datasets, consisting of five distinct species (two human, three rodent) impacted on model performance. Models trained on five-species *Plasmodia* datasets performed just as well as models trained on a single *Plasmodium* dataset related to their specific species (**Figure 4**).

**Figure 4.**
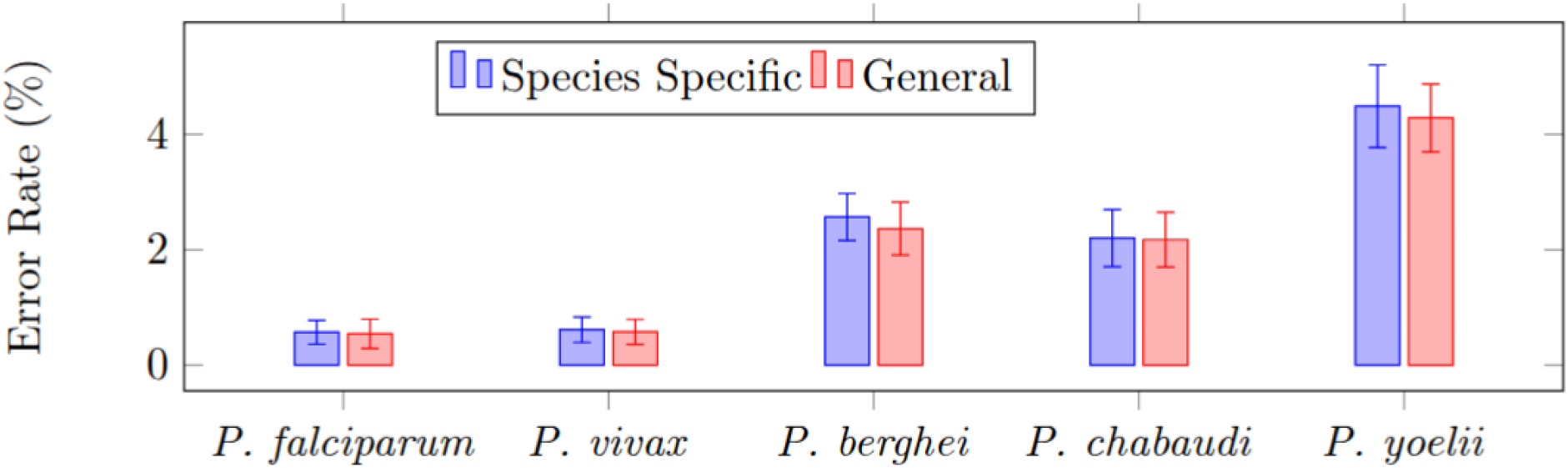
Species Specific dataset vs General dataset Error Rate (percentage of false predictions in set of total predictions). Species specific (blue) are models that were trained solely on the dataset of that species, whereas general (red) are models trained on all five species. The slightly lower error rates from the General model are not statistically significant [P-value 0.215]. All models were evaluated against annotated test data.

This demonstrates that there is no improvement when generating separate models for separate species. A generic all-species model will perform just as well and add greater utility to any platform. When considering the high number of classes (1000 classes in ImageNet-1k) that these image classification models are pre-trained on, it is logical that they can learn the nuance in features across distinct species without losing individual species accuracy. **Figure 5** shows a t-SNE (t-distributed Stochastic Neighbor Embedding) projection of a balanced test set on the final 384-dimensional layer of the classification network (ViT-small). This shows a dimension reduction image of the high-level features present in each cell image with respect to the cell’s predicted infection. Despite the clear clusters that relate to each captured species dataset, there is an obvious learned distinction between infected and uninfected cells across all species (compare round circles with plus symbols). Of note, there is also a significant cluster overlap between the three rodent species (*P. berghei*, *P. chabaudi* and *P. yoelii*), indicating that these datasets, which show similar features, achieve a lower error rate from the general dataset over the species-specific models as any color overlap represents different species expressing the similar visual features.

**Figure 5.**
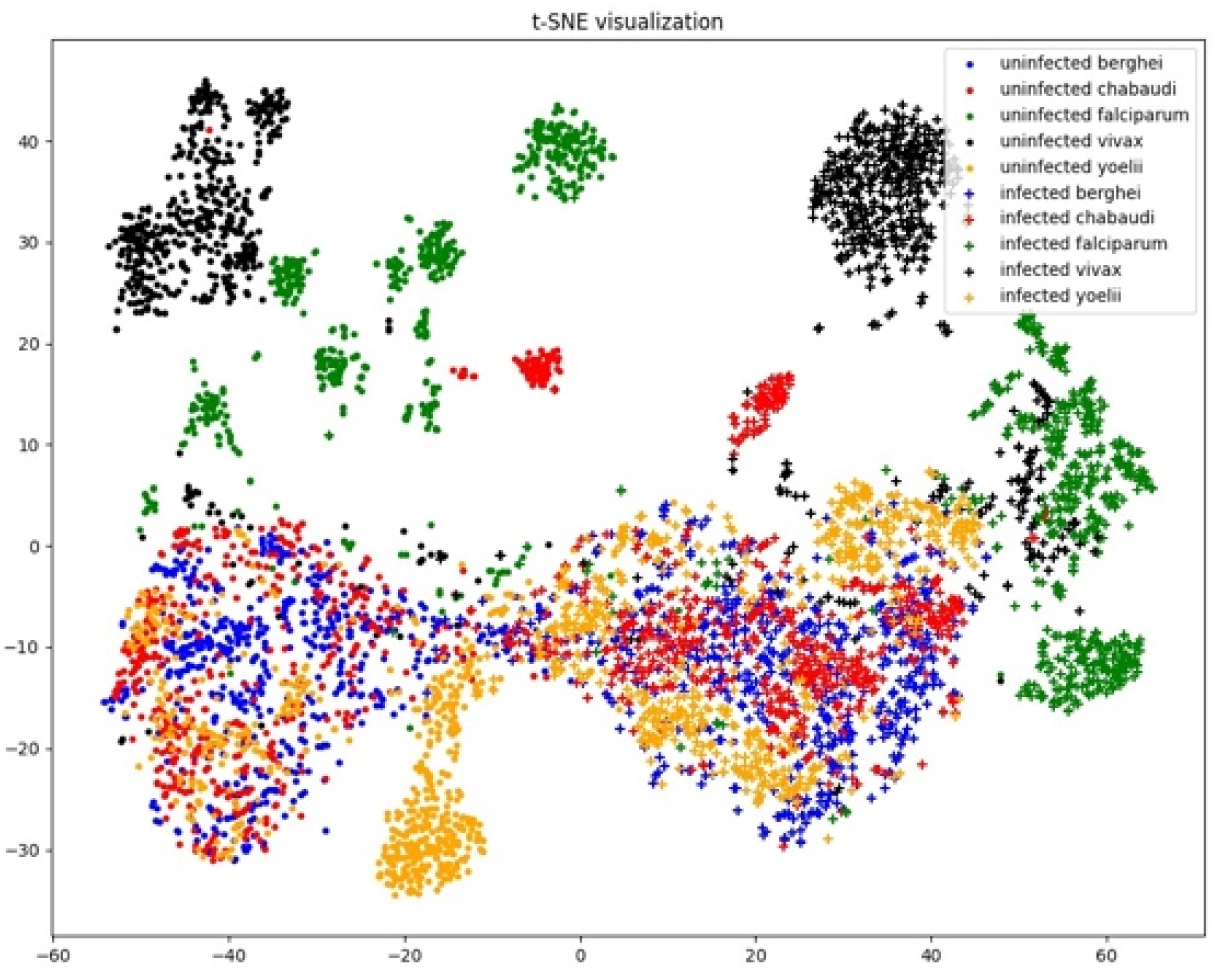
A t-distributed stochastic neighbour embedding (t-SNE) projection of a balanced test set on the final 384-dimensional layer of the classification network (ViT-small). Different colors and shapes represent distinct species and infection status, respectively.

This lower error rate can also be seen subtly in **Figure 4**.

### Fine-tuned Multi-Species Classification Model shows Improved Accuracy with Unseen Species over Single-Species Models

Having demonstrated the ability of models to accurately detect and classify within the distribution of multiple species datasets, we next sought to ask whether the same model could work with parasites of a different “unseen” *Plasmodium* species. To address this, we obtained 128 additional *P. cynomolgi* and *P. knowlesi* images (two species absent in our dataset, a kind gift from T. Annoura & T. Araki, National Institute of Infectious Diseases (NIID), Tokyo, Japan) and passed them through three models trained on three datasets: an all *Plasmodium* species dataset; a *P. falciparum* only dataset; and a *P. vivax* only dataset. The two parasite species, *P. cynomolgi* and *P. knowlesi,* infect wild monkeys but are responsible for a growing, and likely under-reported, prevalence of zoonotic malaria from monkey to human. Phylogenetically, they are more closely related to *P. vivax*^27^ than any of the other malaria species in our combined dataset. Images were passed through the object detection model to obtain 6,668 cell images. Of the cell images, models trained on the three different datasets disagreed on 164 images. **Figure 6** shows the three ROC (Receiver Operating Characteristic) curves comparing the performance of the five species versus single *P. falciparum* and *P. vivax* trained models.

**Figure 6.**
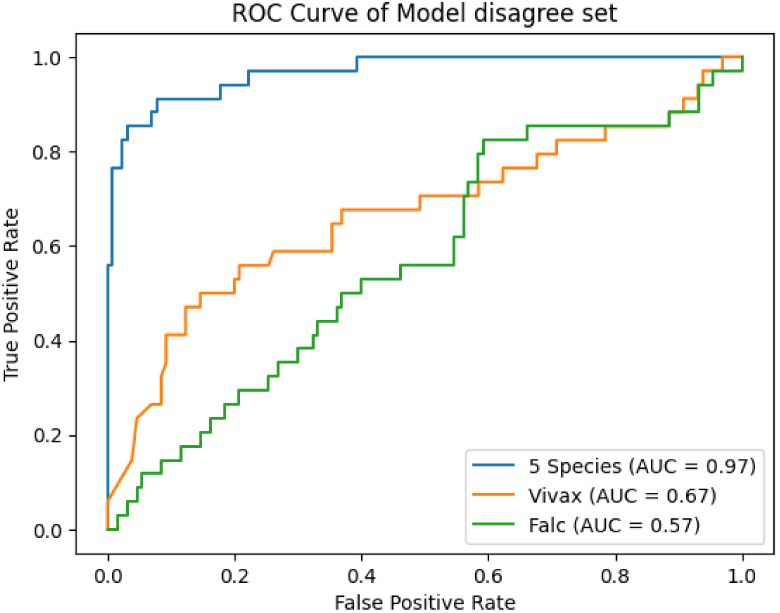
A ROC curve comparison on the set of models’ disagreed cell images of unseen species (*P. cynomolgi* and *P. knowlesi*) data (n=164) between the five species and single species models.

Noticeably, the five species model showed a considerable improvement over the two single-species models. The multi-species dataset performed well in generalising across malaria species, to see data outside of the distribution of the training data, indicating a better model interpretability over the potential distribution of blood-stage malaria parasite phenotypes, including unseen phenotypes. Of note, the phylogenetically more closely related *P. vivax* model performed more accurately than *P. falciparum*, however a DeLong AUC test showed that this result was not significant (p=0.28).

### Fast Requests

Having markedly improved the platform’s ability to detect objects, distinguish infected from uninfected and demonstrated its generalisability to other species, we finally sought to improve the PlasmoCount platform’s overall speed and performance.

Mobile on-device processing eliminates the need for client server interactions, resulting in a significant reduction in request latency. This optimisation is particularly valuable in malaria prevalent regions where there is often low bandwidth. A mobile version of PlasmoCount not only delivers faster, more responsive performance but also offers increased security as a smaller volume of sensitive data is passed over the network. Additionally, on-device processing improves scalability as it heavily reduces the reliance on cloud-based infrastructure.

Figure 7 details the improvements in single image processing time across different iterations of PlasmoCount. Assessing the mobile application versions of the platform, it is clear that these are significantly faster than their predecessors, which should greatly aid microscopic assessment of Giemsa smears. The web version of PlasmoCount stores the image and result files in the cloud so that they are readily available to the end user. The architecture of the PlasmoCount 2.0 web-application streamlines these server storage interactions by batching the requests made to the cloud storage system. Additionally, the server storage time is processed asynchronously meaning that it does not delay the response to the user. PlasmoCount 2.0’s revised model selection with an additional emphasis on speed, shows a significant reduction in inference time with no expense to accuracy. Moreover, the mobile version does not upload image files to a server, and therefore it achieves the fastest time of just under three seconds for one image.

**Figure 7.**
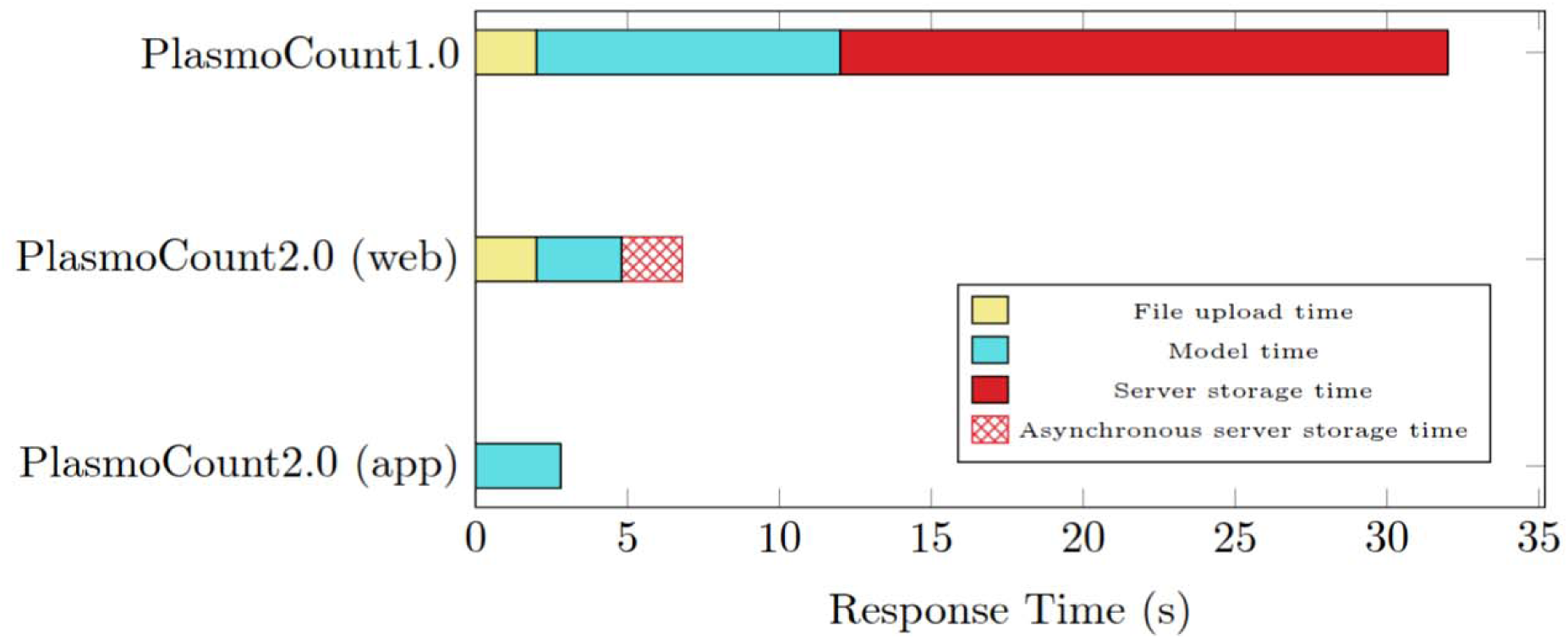
Shows the difference in the time taken to process a single 100x image. ‘PlasmoCount 1.0’ (depreciated) shows the time taken before making any changes, ‘PlasmoCount 2.0’ (depreciated) (web) and ‘PlasmoCount 2.0 ‘(app) show the time taken to the web and app versions after making speed improvements. PlasmoCount 2.0 (app) response time was measured using a Google Pixel 6.

One of the opportunities that arise with routine usage of the PlasmoCount application is the generation of triaged data, over a diversity of microscopy platforms (species, quality, staining & magnification) that has passed some level of expert assessment. This can, in the future, grow the training data set potential, making future improvements in model accuracy possible. Cell editing allows for new images to be incorporated into training data. While various data augmentation techniques can help to mimic this broader distribution, real images from this feature space would allow future models to learn a better representation of any infected cell, avoiding an overfit to any one specific dataset/condition.

## Conclusion

PlasmoCount 2.0 provides its users with a fast and interactive application. It improves upon the original PlasmoCount 1.0 platform by bolstering inference speed and providing domain generalisation to the model pipeline in the form of multi-magnification and multi-species augmentations. Moreover, transitioning from a client-server architecture to a mobile platform not only improves processing speed but also provides greater robustness and reliability in low-connectivity environments.

Further work focusing on analysing and improving PlasmoCount’s diagnostic performance would open the tool up to a wider audience both as a lab tool and potentially a clinical aid in diagnostics labs where imaging is still routinely used. Integrating leukocyte prediction into the model pipeline would be invaluable for this latter context, as these cells are scarce in the current dataset.

PlasmoCount 2.0 currently includes multiple new features that increase its scope considerably. Multi-species and multi-magnification inference drastically increase the dimensionality of input images that the application can reliably predict on, bolstering the accuracy on out of distribution images and thus making it a viable tool for a wider user base. We have also provided improvements through reducing request latency, improving the average user response time by over 90%. We achieved this by introducing an optimized storage upload architecture, selecting the optimal neural networks for each task in the model pipeline. Further enhancements to the frontend bolster the application’s usability. PlasmoCount 2.0 provides a practical application that utilizes all the benefits of light microscopy for parasite counting, whilst ameliorating the time-consuming process of manual counting. Given these improvements, we expect that PlasmoCount 2.0 will prove a valuable tool for researchers and clinicians in the detection and counting of malaria parasite infected cells.

## Data Availability

All data produced in the present study are available upon reasonable request to the authors.

## Acknowledgement

The authors would like to thank A. Cunnington and A. Georgiadou (Imperial College London, UK), S. Reece and A. O’Donnell (University of Edinburgh, UK), F. Firschknecht (University of Heidelberg, Germany), J. Langhorne (Francis Crick Institute, UK) and T. Annoura and T Araki (NIID, Tokyo, Japan) for images of rodent and non-human primate malaria Giemsa smears. We thank Mira Davidson and members of the Baum lab for their helpful comments in reviewing the manuscript. JB acknowledges previous funding from Wellcome UK (Investigator 100993/Z/13/Z) and RoseTrees Trust UK (PGS21/10035) as well as current funding from the National Health and Medical Research Council (NHMRC) of Australia (APP2026574). All data produced in the present study are available upon reasonable request to the authors.

## Author contributions

Conceptualization: FW, JB Methodology: FW, YL, DN Investigation: FW, YL, DN Supervision: JB, EM Writing—original draft: FW, JB Writing—review & editing: All Authors

## References

1. WHO, World Malaria Report 2024. (WHO, 2024).

2. Feleke, S. M. et al. Plasmodium falciparum is evolving to escape malaria rapid diagnostic tests in Ethiopia. Nature Microbiology 6, 1289–1299 (2021).

3. Davidson, M. S. et al. Automated detection and staging of malaria parasites from cytological smears using convolutional neural networks. Biological Imaging 1, (2021).

4. Halim, S., Bretschneider, T. R., Li, Y., Preiser, P. R. & Kuss, C. Estimating Malaria Parasitaemia from Blood Smear Images. in 1–6 (2006). doi:10.1109/ICARCV.2006.345381.

5. Purwar, Y., Shah, S. L., Clarke, G., Almugairi, A. & Muehlenbachs, A. Automated and unsupervised detection of malarial parasites in microscopic images. Malaria Journal 10, 364– 364 (2011).

6. Anggraini, D. et al. Automated status identification of microscopic images obtained from malaria thin blood smears using bayes decision: A study case in plasmodium falciparum. in 347–352 (2011).

7. Di Ruberto, C., Dempster, A., Khan, S. & Jarra, B. Analysis of infected blood cell images using morphological operators. Image and Vision Computing 20, 133–146 (2002).

8. Poostchi, M., Silamut, K., Maude, R. J., Jaeger, S. & Thoma, G. Image analysis and machine learning for detecting malaria. Translational Research 194, 36–55 (2018).

9. Rajaraman, S. et al. Pre-trained convolutional neural networks as feature extractors toward improved malaria parasite detection in thin blood smear images. PeerJ 6, (2018).

10. Hung, J. & Carpenter, A. E. Applying Faster R-CNN for Object Detection on Malaria Images. Conf Comput Vis Pattern Recognit Workshops (2017) doi:10.1109/cvprw.2017.112.

11. Dong, Y. et al. Evaluations of deep convolutional neural networks for automatic identification of malaria infected cells. in 101–104 (2017). doi:10.1109/BHI.2017.7897215.

12. Liang, Z. et al. CNN-based image analysis for malaria diagnosis. in 493–496 (2016). doi:10.1109/BIBM.2016.7822567.

13. Umer, M. et al. A Novel Stacked CNN for Malarial Parasite Detection in Thin Blood Smear Images. IEEE Access 8, 93782–93792 (2020).

14. Najman, L. & Schmitt, M. Watershed of a continuous function. Signal Processing 38, 99–112 (1994).

15. Liu, R. et al. AIDMAN: An AI-based object detection system for malaria diagnosis from smartphone thin-blood-smear images. Patterns 4, 100806 (2023).

16. Tan, D. & Liang, X. Multiclass malaria parasite recognition based on transformer models and a generative adversarial network. Scientific Reports 13, (2023).

17. Thomas, K., Tchiotsop, D., Tonye, E., Ele, P. & Belong, E. Automated Diagnosis of Malaria in Tropical Areas Using 40X Microscopic Images of Blood Smears. International Journal of Biometrics and Bioinformatics 10, 12–23 (2016).

18. Yanik, S. et al. Application of Machine Learning in a Rodent Malaria Model for Rapid, Accurate, and Consistent Parasite Counts. The American Journal of Tropical Medicine and Hygiene (2024) doi:10.4269/ajtmh.24-0135.

19. Yu, H. et al. Malaria Screener: a smartphone application for automated malaria screening. BMC Infectious Diseases 20, (2020).

20. Hung, J. et al. Keras R-CNN: library for cell detection in biological images using deep neural networks. BMC bioinformatics 21, 300 (2020).

21. Kassim, Y. et al. Clustering-Based Dual Deep Learning Architecture for Detecting Red Blood Cells in Malaria Diagnostic Smears. IEEE Journal of Biomedical and Health Informatics **PP**, 1– 1 (2020).

22. Carion, N. et al. End-to-End Object Detection with Transformers. (2020).

23. Deng, J. et al. ImageNet: A large-scale hierarchical image database. in 248–255 (2009). doi:10.1109/CVPR.2009.5206848.

24. Tan, H. H. & Lim, K. H. Vanishing Gradient Mitigation with Deep Learning Neural Network Optimization. in 1–4 (2019). doi:10.1109/ICSCC.2019.8843652.

25. Dosovitskiy, A. et al. An Image is Worth 16x16 Words: Transformers for Image Recognition at Scale. Preprint at http://arxiv.org/abs/2010.11929 (2021).

26. Liu, Z. et al. A ConvNet for the 2020s. in 11976–11986 (2022).

27. Galen, S. C., et al. (2018). The polyphyly of *Plasmodium*: comprehensive phylogenetic analyses of the malaria parasites (order Haemosporida) reveal widespread taxonomic conflict. Royal Society Open Science, 5(5), 171780. doi:10.1098/rsos.171780

